# Increased serum levels of soluble TNF-α receptor is associated with mortality of ICU COVID-19 patients

**DOI:** 10.1101/2020.07.12.20152066

**Authors:** Esmaeil Mortaz, Payam Tabarsi, Hamidreza Jamaati, Neda Dalil Roofchayee, Neda K.Dezfuli, Seyed Mohamadreza Hashemian, Afshin Moniri, Majid Marjani, Majid Malkmohammad, Davood Mansouri, Mohammad Varahram, Gert Folkerts, Ian M Adcock

## Abstract

**Background:** Severe acute respiratory syndrome coronavirus 2 (SARS-CoV-2) that causes coronavirus disease 2019 (COVID-19) has spread to almost 100 countries, infected over 10M patients and resulted in 505K deaths worldwide as of 30^th^ June 2020. The major clinical feature of severe COVID-19 requiring ventilation is acute Respiratory Distress Syndrome (ARDS) with multi-functional failure as a result of a cytokine storm with increased serum levels of cytokines such as TNF-α and IL-6 being reported. TNF-α levels are increased during the cytokine storm in very ill patients and soluble receptors for IL-6 and IL-2 are present in the blood of COVID-19 patients,

**Objectives:** To elucidate the involvement of serum levels of soluble TNF-Receptor of severe and mild COVID-19 patients to determine for severity of disease.

**Method:** We recruited16 severe COVID-19 patients in the ICU on ventilator support and 26 milder COVID-19 patients who were hospitalised but not within the intensive care unit (ICU) between March-May 2020 at the Masih Daneshvari Hospital Tehran, Iran. After harvesting of whole blood the serum was isolated and soluble TNF-Receptor levels measured by ELISA.

**Results:** Serum levels of the usually inhibitory soluble TNF-α receptor 1 (sTNFαR1) were significantly elevated in severe COVID-19 patients at admission to ICU. High serum levels of sTNFαR1 were associated with mortality of severe COVID-19 patients treated within ICU.

**Conclusions:** This pilot study demonstrates for role of STNF-αR1 receptor in severity of disease. Future studies should examine whether lower levels of systemic sTNFαR1 at admission may indicate a better disease outcome.

## Introduction

Understanding the mechanism(s) and profiles of the cytokine storm observed in Coronavirus disease 2019 (COVID-19) is crucial for the development of effective therapeutic interventions. As such, cytokine blockers and immune-host modulators are currently being tested in severely ill COVID-19 patients to cope with the overwhelming systemic inflammation (1). Most current reports highlight different elevated cytokine patterns among severely ill COVID-19 patients that further indicates the potential for already existing immunosuppressive agents in COVID-19.

COVID-19 is an infectious disease caused by severe acute respiratory syndrome coronavirus 2 (SARS-CoV-2) with symptoms such as fever, dry cough and shortness of breath (2). According to the disease state, patients are divided into two major groups; asymptomatic or mild cases that usually recover after most showing only mild to moderate symptoms. However, severe cases (approximately 15%) develop multi-organ failure due to a cytokine storm resulting in respiratory failure that requires intensive care unit (ICU) admission (3, 4).

COVID-19 patients that need ICU admission have higher blood concentrations of IL-6, CXCL10, CCL2 and TNF-α as compared to milder patients that do not require ICU admission

Most inflammatory mediators in severe COVID-19 patients are probably produced by immune effector cells that are able to release large amounts of pro-inflammatory cytokines that contribute to the sustained systemic inflammation (5).

A pathogenic role for IL-6 and its receptor in COVID-19 has been described (6,7).

In contrast, a negative relationship between the concentration of soluble IL-2 receptor (sIL-2R) and T-cell number in the blood from COVID-19 patients has recently has been reported.(8). TNF-α is produced primarily as a transmembrane precursor by monocytes/macrophages, but a number of other cell types, such as T and B lymphocytes, mast cells, natural killer cells, neutrophils, fibroblasts, and osteoclasts, also secrete this cytokine (9). The TNF-α precursor is cleaved by TNFα-converting enzyme (TACE) to liberate soluble TNF (sTNF) which circulates in blood to enable its effect on distant sites. Upon release, TNF binds to two distinct membrane receptors on target cells: TNFR1 and TNFR2 (10). TNFR1 is ubiquitously expressed within the lymphoid system and nearly all cells of the body, which likely accounts for TNF’s wide-ranging functions. TNFR2 expression is limited to certain lymphocyte populations including T-regulatory cells (T_regs_) (11). Generally, TNF depends on TNFR1 for apoptosis due to the presence of death domains and TNFR2 for any function related to cell survival, although there is some degree of overlap depending upon the activation state of the cell and other factor (12).

The role of these soluble forms of TNFR1 and TNFR2 is debated as they bind to and inhibit TNF-α function during acute inflammation. In contrast, sTNF-α/sTNFR1 complexes can act to enhance TNF-α function under chronic inflammatory conditions by slowly releasing TNF-α (13). However, these effects may also be concentration-dependent. In previous studies, circulating levels of sTNFR1 is associated with adverse outcomes and severity of myocardial infarction, mortality in subjects with a ruptured abdominal aortic aneurysm, the risk of death and of death in patients with diabetic nephropathy and to adverse outcomes in diabetic patients (14).

## Materials and Methods

In this study we measured the serum levels of sTNF-α in COVID-19 patients of different severity. 16 severe COVID-19 patients in the ICU on ventilator support and 26 milder COVID-19 patients who were hospitalised but not within the ICU were recruited between March-May 2020 at the Masih Daneshvari Hospital Tehran, Iran. All patients were RT-PCR positive for SARS-CoV-2. Moreover, 7 non-COVID-19 healthy age- and gender-matched control subjects were enrolled in this study. The demographic data of patients are provided in Table 1. Serum was obtained from peripheral blood of subjects and ELISA used to detect the expression of Human Soluble TNF Receptor I (ab100642) (Cambridge, United Kingdom).

**Table 1.**
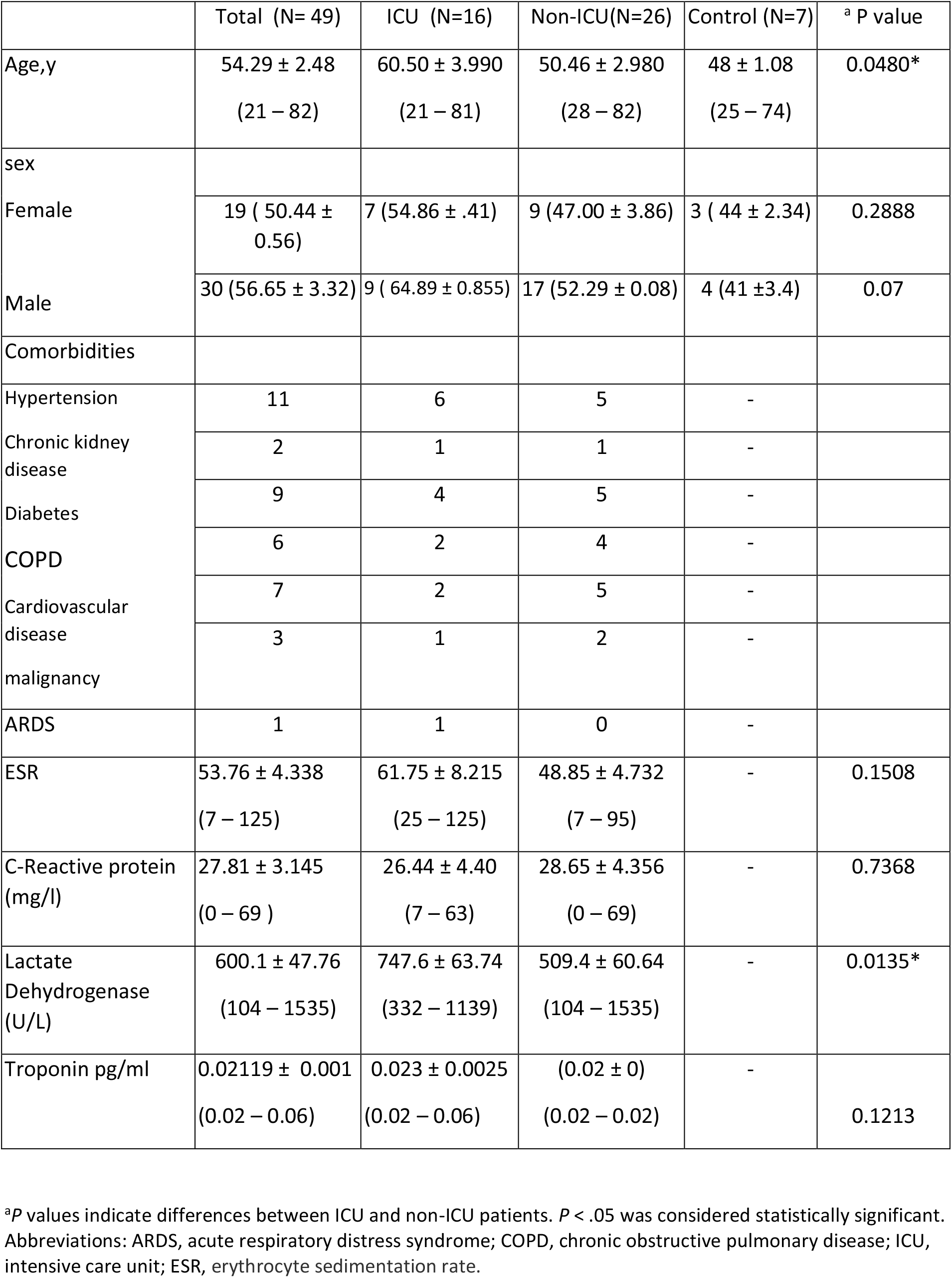
Demographic information of severe (ICU) and mild (Non-ICU) COVID-19 inpatients.

## Results

Severe ICU COVID-19 patients showed significantly increased levels of sTNF-α (P≤0.05, Fig 1). In contrast, the expression of sTNF-αR1 in mild COVID-19 patients was lower than in healthy control subjects although this did not reach significance. This data suggests that there is an association between the severity and high serum levels of sTNF-αR1 which may contribute to multifunctional failure in COVID-19 patients. Most (80%) severe patients died whereas all mild COVID-19 patients survived and were discharged from the hospital.

**Fig. 1.**
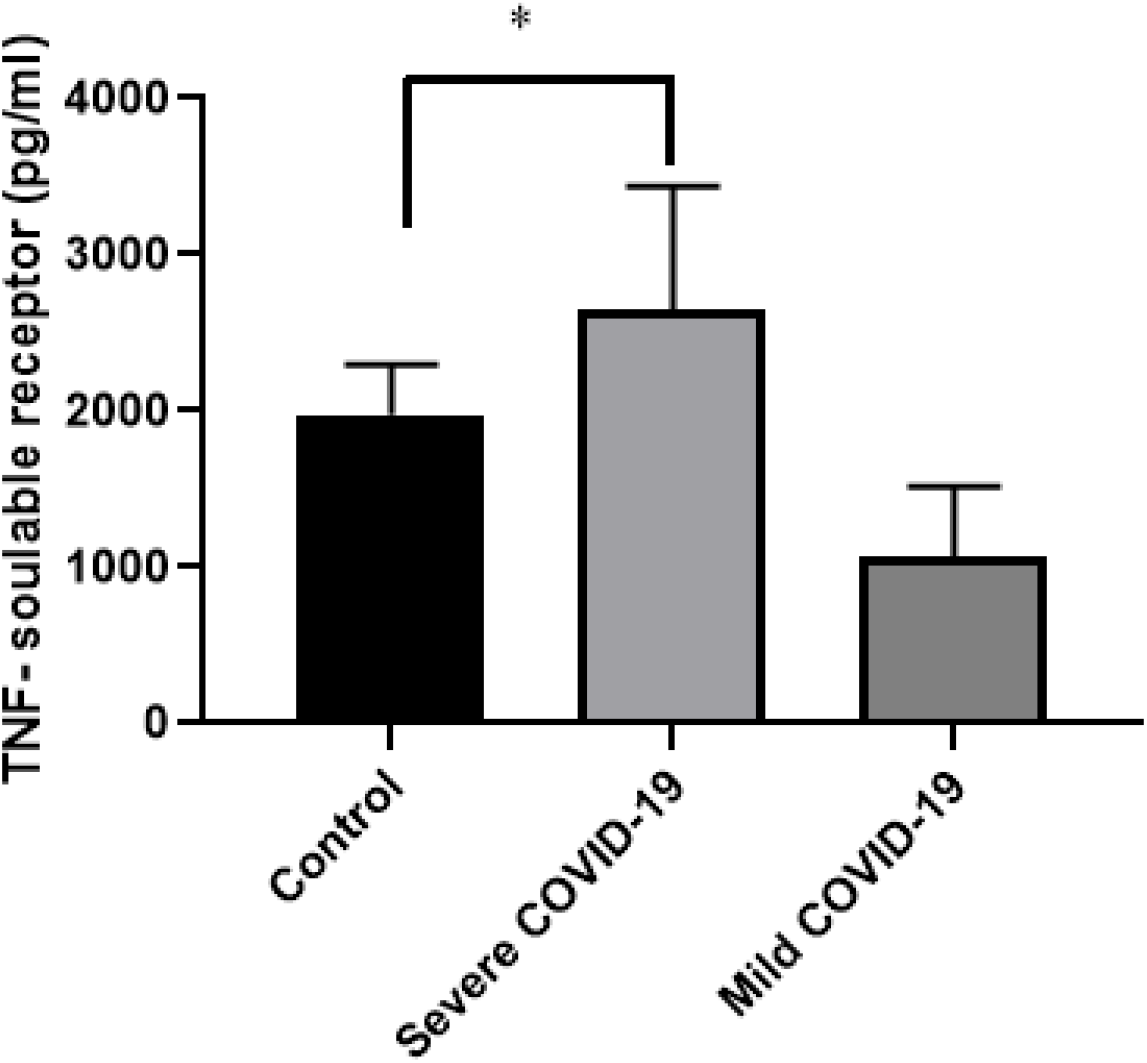
The serum levels of soluble TNF receptors in severe, mild COVID-19 patients and control subjects. The serum was subjected to ELISA methods as described in material and methods.. Asterisks indicated to statistically significance between controls and sever COVID-19 patients (P≤0.05).

## Discussion

A cytokine storm with uncontrolled raised levels of inflammatory mediators has been reported in COVID-19 infected patients. As such, elevated levels of some inflammatory cytokines have been proposed as predictors of survival and of severity in patients treated within ICU (7) In contrast, some severe COVID-19 subjects express low blood cytokine levels due to immune cell exhaustion (15-18).

Elucidation of a possible predictor of COVID-19 severity and survival of patients at admission to ICU is important as this will affect treatment options. In this pilot study we investigated the levels of STNF-αR1 in the serum of ICU- and non-ICU-treated COVID-19 patients. We found a significant association between disease severity and with serum levels of sTNFαR1.

To our knowledge this is the first report of increased serum levels of sTNFαR1 in severe COVID-19 patients. Most importantly, our data indicates that high serum levels of sTNFαR1 are associated with mortality of COVID-19 patients within ICU. Future studies should examine whether lower levels of systemic sTNFαR1 at admission may indicate a better disease outcome.

## Data Availability

all data included in the paper

## Footnotes

## Acknowledgement

We are acknowledge all participates whom are alive and remember those patients in this study who died of COVID-19.

## Statement of Ethics

The study was approved by Ethical committee of Masih Daneshvari Hospital IR.SBMU.NRITLD.REC.1399.123.

## Author Contributions

EM, NDR, NKD did experiments. EM wrote the draft of paper.PT, HRJ, SMH, MM, MMR and AM provided the patients and samples. DM and IMA revised the paper. IMA approved the final version as corresponding author.

